# The pattern of antidiabetic drugs and glycaemic control among type 2 diabetes patients in an Endocrinology Clinic in Lagos, Nigeria

**DOI:** 10.1101/2023.06.25.23291774

**Authors:** Taoreed Adegoke Azeez

## Abstract

**Background:** Diabetes mellitus is highly prevalent in Nigeria. In addition to lifestyle changes, hypoglycaemic agents are of crucial importance in providing optimal care. The study aimed to study the pattern of hypoglycaemic agents and glycaemic control.

**Methods:** It is a retrospective study. Parameters of interest were obtained from the electronic medical records of 248 type 2 diabetes patients.

**Results:** The mean age of the patients was 59.6 ± 12.7 years. Biguanides (85.5%) and DPP-IV inhibitors (60.9%) were the most commonly used antidiabetic drugs, while thiazolidinediones (1.2%), α-glucosidase inhibitors (0.8%), and meglitinides (0%) were the least prescribed. In addition, SGLT-2 inhibitors, insulins, GLP-1 agonists, and sulphonylureas were prescribed to 31.0%, 17.1%, 15.3%, and 14.9%, respectively. On average, about 2-3 hypoglycaemic agents were prescribed. The number of drugs used and HbA1c were significantly negatively correlated. Biguanide, insulins, and DPP-IV inhibitors were associated with a significantly lower HbA1c.

**Conclusions:** Metformin is the drug of choice. Combining 2 to 3 drugs is very common. Glycaemic control is better in the present study, compared with prior studies, and it could be due to the pattern of drugs used in this study.

## Background

Diabetes mellitus is a common metabolic disorder in Nigeria, and its prevalence was reported at 5.8%.^1^ The prevalence is even higher in urban areas such as Lagos.^2^ Type 2 diabetes is the most common form of diabetes. The management of type 2 diabetes often requires lifestyle changes as well as the use of medications. These medications have different mechanisms of action with variable efficacy. The pharmacologic treatment aims to optimize glycaemic control, reduce the risk of complications or even death, and promote a good quality of living.

The classes of antidiabetic drugs, sometimes described as hypoglycaemic agents, used in the management of type 2 diabetes include sulphonylureas (e.g., gliclazide and glimepiride), biguanides (e.g. metformin), dipeptidyl peptidase IV (DPP-IV)-inhibitors (e.g., vildagliptin and sitagliptin), glucagon-like peptide-1 (GLP-1) agonists (e.g., liraglutide and dulaglutide), and sodium-glucose cotransporter 2 (SGLT-2) inhibitors (e,g., dapaglifozin and empaglifozin). Other classes are alpha-glucosidase inhibitors (e.g., acarbose and voglibose), amylin mimetics (e,g., pramlintide), peroxisome proliferator-activated receptor-gamma (PPAR-□) agonists, otherwise known as thiazolidinediones (e.g., pioglitazone), and meglitinides (e.g., repaglinide and nateglinide). Insulins (basal, pre-mixed, or basal-bolus) are sometimes used in the management of type 2 diabetes. The factors that determine the choice of medications include efficacy, co-morbidities, risk of hypoglycaemia, cardiovascular safety, weight gain, potential side effects, and cost.^3^

Glycated haemoglobin (HbA1c) is a widely validated metric for assessing the degree of glycaemic control.^4^ HbA1c reflects the glycaemic profile in the preceding 2-3 months. The American Diabetic Association (ADA) recommends an HbA1c of less than 7% for non-pregnant adults without significant hypoglycaemia.^5^ However, the American Association of Clinical Endocrinologists/American College of Endocrinology (AACE/ACE) recommended an HbA1c of less than 6.5%, provided that it can be achieved in a safe and affordable manner.^6^ There is a paucity of data on the proportion of diabetic patients with good glycaemic control in a private tertiary setting in an urban centre, where the cost of care is typically affordable.

The study aimed to determine the pattern of antidiabetic drugs used by patients with type 2 diabetes and assess their degree of glycaemic control.

## Methods

It was a retrospective study of patients with type 2 diabetes who attended the outpatient department of the Reddington Multi-Specialist Hospital, Lagos, Nigeria. The study lasted from 1^st^ of June 2021 to 31^st^ of May, 2023. Reddington Multi-Specialist Hospital is one of the biggest private tertiary hospitals in West Africa, and it attends to various patients from the sub-region. The total sample size was 248. The study population involved adults who were above 18 years and had attended the out-patient department for at least 6 months. Patients with type 1 diabetes and gestational diabetes were excluded from the study. Individuals with malignancies and pregnant women were similarly excluded. In addition, anyone who had been managed for a diabetic emergency in the preceding 3 months or who had incomplete information in the electronic records was excluded from the study.

All the relevant information was obtained from the electronic medical records of the respective patients. Information obtained included the age, gender, modality of payment, systolic blood pressure (SBP), diastolic blood pressure (DBP), weight, height, and current antidiabetic drugs of the patients. The body mass index (BMI) was calculated using the formula below.^7^ BMI is classified using the World Health Organization (WHO) categories, as illustrated in *table 1*.^8^ BMI=Weight (kg)/Height (m)^2^

**Table 1:**
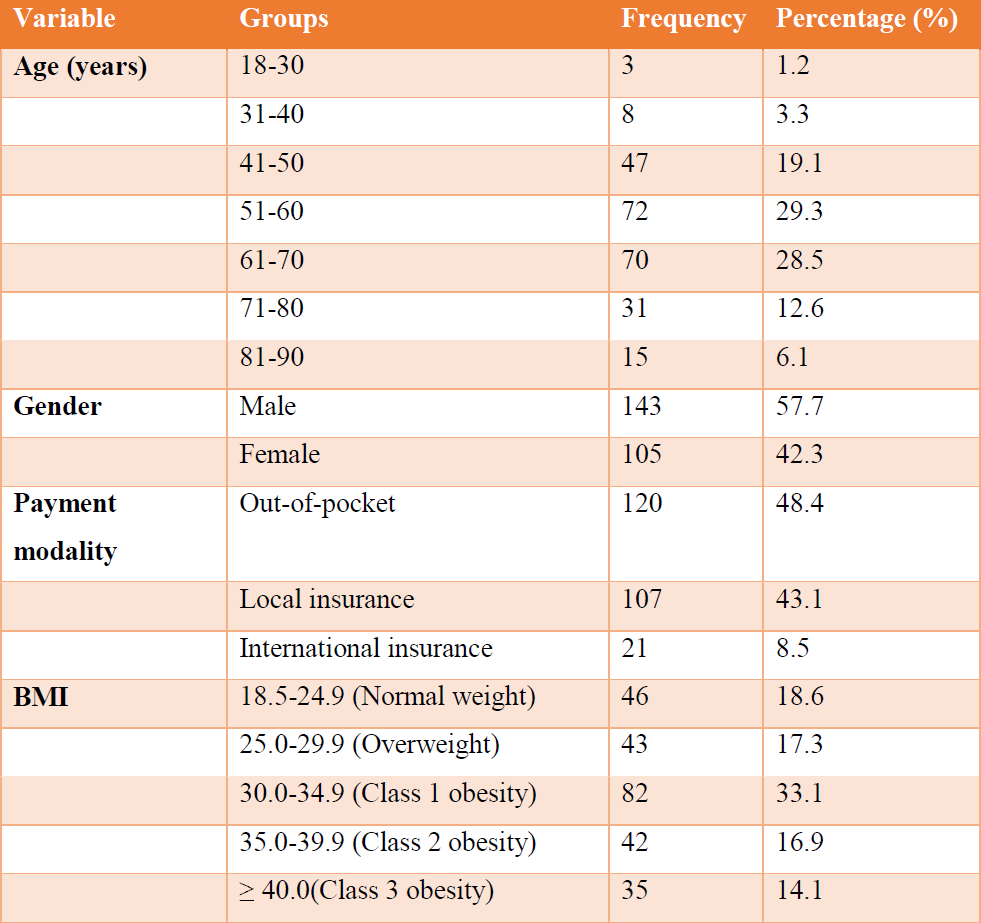
Characteristics of the participants

| **Variable** | **Groups** | **Frequency** | **Percentage (%)** |
| --- | --- | --- | --- |
| **Age (years)** | 18-30 | 3 | 1.2 |
|  | 31-40 | 8 | 3.3 |
|  | 41-50 | 47 | 19.1 |
|  | 51-60 | 72 | 29.3 |
|  | 61-70 | 70 | 28.5 |
|  | 71-80 | 31 | 12.6 |
|  | 81-90 | 15 | 6.1 |
| **Gender** | Male | 143 | 57.7 |
|  | Female | 105 | 42.3 |
| **Payment modality** | Out-of-pocket | 120 | 48.4 |
|  | Local insurance | 107 | 43.1 |
|  | International insurance | 21 | 8.5 |
| **BMI** | 18.5-24.9 (Normal weight) | 46 | 18.6 |
|  | 25.0-29.9 (Overweight) | 43 | 17.3 |
|  | 30.0-34.9 (Class 1 obesity) | 82 | 33.1 |
|  | 35.0-39.9 (Class 2 obesity) | 42 | 16.9 |
|  | ≥ 40.0(Class 3 obesity) | 35 | 14.1 |

Ethical approval was obtained from the Ethical Committee of the Hospital group. The data was initially meticulously collected on a Microsoft Excel sheet and subsequently analyzed using the Social Science Statistical Package (SPSS) version 26. The data were presented in tables and charts. Chi-square test was used to test the association between categorical variables, while Pearson’s correlation was used to test the relationship between continuous variables. The Student’s T-test was used to compare means between two continuous variables. A p-value below 0.05 was considered statistically significant.

## Results

The study involved 248 individuals. The average age was 59.6 ± 12.7 years. The majority of the participants (57.8%) are between 40-60 years of age. The mean BMI was 32.4±7.4 kg/m^2^. The mean systolic blood pressure was 132.6±15.1 while the mean diastolic blood pressure was 77.8±11.2. The majority of the participants (89.4%) had a blood pressure of less than 140/90 mmHg. *Table 1* illustrates the other characteristics of the participants. The total number of participants paid for by various insurance modalities (51.6%) were slightly more than those paying out-of-pocket (48.4%). About two-thirds of the participants were obese.

The classes of antidiabetic drugs being used are depicted in *figure 1*. In this study, biguanides (85.5%) and DPP-IV inhibitors (60.9%) were the most commonly used antidiabetic drugs. Insulin was used by 17.3% of the participants. About 2.4% of the participants were being managed with lifestyle therapy only. The frequency of usage of individual antidiabetic drugs is illustrated in *table 2*. Metformin (85.5%) and Vildagliptin (56.5%) were the most frequently prescribed antidiabetic drugs, according to this study. *Table 3* depicts how the drugs tended to be combined among the patients. As shown in *figure 2*, 28% and 35% of the participants took 2 and 3 antidiabetic drugs, respectively.

**Figure 1:**
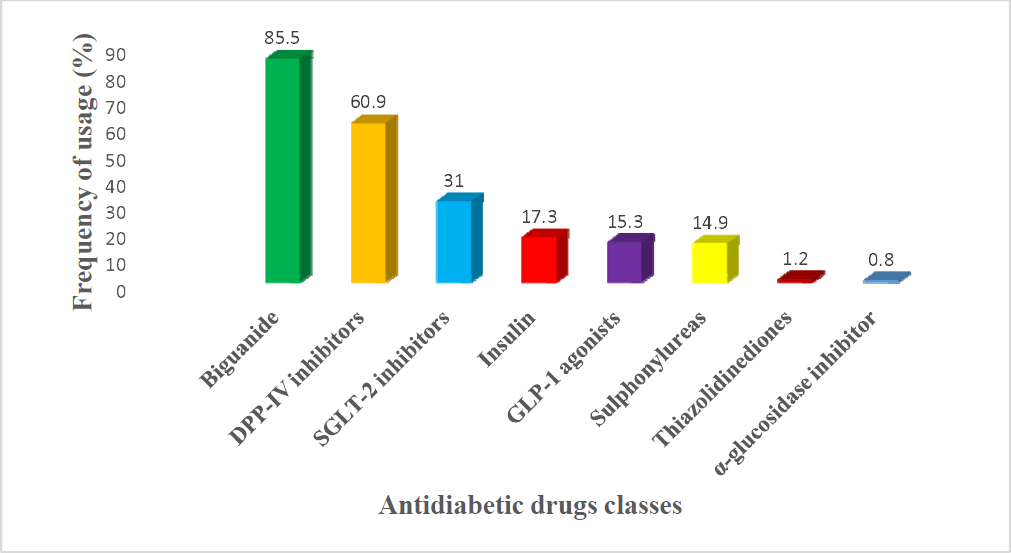
Classes of antidiabetic drugs and their frequency of usage

**Table 2:** Frequency of usage of the various antidiabetic drugs

| Class | Specific drugs | Frequency | Percentage (%) |
| --- | --- | --- | --- |
| Biguanides | Metformin | 212 | 85.5 |
| DPP-IV inhibitors | Vildagliptin | 140 | 56.5 |
|  | Linagliptin | 8 | 3.2 |
|  | Sitagliptin | 3 | 1.2 |
| SGLT-2 inhibitors | Dapaglifozin | 73 | 29.4 |
|  | Empaglifozin | 4 | 1.6 |
| Insulins | Glargine | 39 | 15.7 |
|  | Degludec | 3 | 1.2 |
|  | Aspart | 3 | 1.2 |
| GLP-1 agonists | Dulaglutide | 20 | 8.1 |
|  | Liraglutide | 16 | 6.5 |
|  | Semaglutide | 3 | 1.2 |
| Sulphonylureas | Gliclazide | 27 | 10.9 |
|  | Glimepiride | 8 | 3.2 |
|  | Glibenclamide | 3 | 1.2 |
| Thiazolidinediones | Pioglitazone | 3 | 1.2 |
| α-glucosidase inhibitor | Voglibose | 2 | 0.8 |

**Table 3:** Combinations of classes of antidiabetic drugs

| **1 drug** | Frequency | Percentage (%) |
| --- | --- | --- |
| Biguanide | 39 | 83 |
| DPP-IV inhibitor | 3 | 6.4 |
| Insulins | 3 | 6.4 |
| SGLT-2 inhibitor | 1 | 2.1 |
| Sulphonylureas | 1 | 2.1 |
| Total | 47 | 100 |
| **2 drugs** | Frequency | Percentage (%) |
| Biguanide + DPP-IV inhibitor | 49 | 57.6 |
| Biguanide + SGLT-2 inhibitor | 13 | 15.3 |
| Biguanide + GLP-1 agonists | 4 | 4.7 |
| DPP-IV inhibitor + Insulins | 4 | 4.7 |
| SGLT-2 + GLP-1 agonists | 4 | 4.7 |
| Biguanide + Sulphonylurea | 3 | 3.5 |
| Biguanide + Insulin | 2 | 2.4 |
| DPP-IV inhibitor + SGLT-2 inhibitor | 2 | 2.4 |
| DPP-IV inhibitor + Sulphonylurea | 2 | 2.4 |
| Insulin + Sulphonylurea | 1 | 1.2 |
| SGLT2 + Insulins | 1 | 1.2 |
| Total | 85 | 100 |
| **3 drugs** | Frequency | Percentage (%) |
| Biguanide + DPP-IV inhibitor + SGLT-2 inhibitor | 23 | 42.6 |
| Biguanide + DPP-IV inhibitor + Sulphonylurea | 14 | 25.9 |
| Biguanide + DPP-IV inhibitor + GLP-1 agonist | 4 | 7.4 |
| Biguanide + DPP-IV inhibitor + GLP-1 agonist | 3 | 5.6 |
| Biguanide + Insulin + GLP-1 agonist | 3 | 5.6 |
| Biguanide + Sulphonylurea + Thiazolidinedione | 1 | 1.9 |
| SGLT-2 inhibitors + GLP-1 agonist + Sulphonylurea | 1 | 1.9 |
| Biguanide + Sulphonylurea + insulin | 1 | 1.9 |
| DPP-IV inhibitor +insulin + Sulphonylurea | 1 | 1.9 |
| Biguanide + SGLT-2 inhibitor + GLP-1 agonist | 1 | 1.9 |
| DPP-IV inhibitor + SGLT-2 inhibitor + GLP-1 agonist | 1 | 1.9 |
| DPP-IV inhibitor + SGLT-2 inhibitor + Sulphonylurea | 1 | 1.9 |
| Total | 54 | 100 |
| **4 drugs** | Frequency | Percentage (%) |
| Biguanide + DPP-IV inhibitors + SGLT-2 inhibitor + insulin | 19 | 57.6 |
| Biguanide + DPP-IV inhibitors + SGLT-2 inhibitor + GLP-1 agonist | 4 | 12.1 |
| Biguanide + DPP-IV inhibitor + SGLT-2 inhibitor + Sulphonylurea | 3 | 9.1 |
| Biguanide + DPP-IV inhibitor + insulin + Sulphonylurea | 2 | 6.1 |
| Biguanide + DPP-IV inhibitor + GLP-1 agonists + insulin | 1 | 3 |
| DPP-IV inhibitor + Insulin + Sulphonylurea + Thiazolidinedione | 1 | 3 |
| Biguanide + insulin + SGLT-2 inhibitor + GLP-1 agonist | 1 | 3 |
| Biguanide + DPP-IV inhibitor + Insulin + Thiazolidinedione | 1 | 3 |
| Biguanide + SGLT-2 inhibitor + Insulin + α-glucosidase inhibitors | 1 | 3 |
| Total | 33 | 100 |
| **5 drugs** | Frequency | Percentage (%) |
| Biguanide + DPP-IV inhibitor + SGLT-2 inhibitor + GLP-1 agonist + Insulin | 4 | 100 |

**Figure 2:**
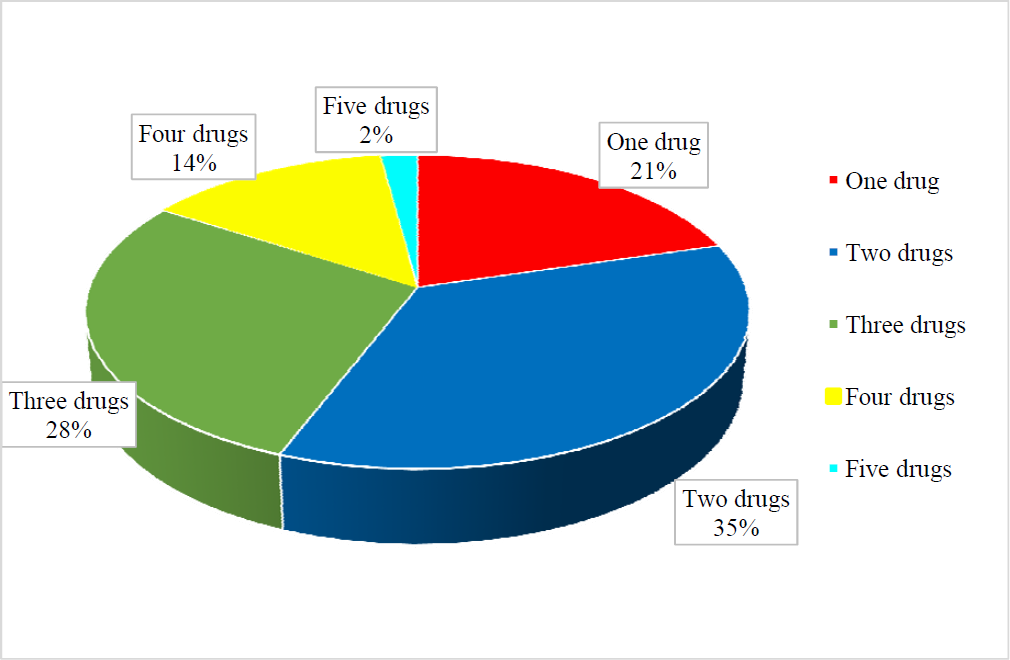
Number of antidiabetic drugs

There was a statistically significant association between the number of antidiabetic drugs used and the age groups (Pearson Chi-square =45.06, p=0.038). In addition, there was a statistically significant association between the number of antidiabetic drugs used and the BMI categories (Pearson Chi-square =30.04, p=0.044). However, there was no statistically significant association between the modality of payment and the number of antidiabetic drugs prescribed to the participants (Pearson Chi-square =6.01, p=0.814).

In terms of glycaemic control, the mean HbA1c of the participants was 7.2±1.99 %. The proportion of participants with optimal glycaemic control, using the ADA and AACE guidelines, is shown in *table 4*. As expected, there was a significant correlation between HbA1c and the number of antidiabetic drugs used (Pearson correlation coefficient= −0.41,p=0.00). However, there was no statistically significant association between the modality of payment and the degree of glycaemic control (Pearson chi-square=1.57,p=0.46). *Table 5* shows the differences in the means, using the T-test, of HbA1c, BMI, SBP and DBP across the different antidiabetic classes of drugs. The use of biguanide, insulins and DPP-IV inhibitors significantly lowered HbA1c. Also, the use of biguanide resulted in a statistically significant lower DBP. Interestingly, in this study, there were no statistically significant differences in the BMI of the patients regardless of whether insulin, SGLT-2 inhibitors, GLP-1 agonists, or sulphonylureas were being used or not.

**Table 4:**
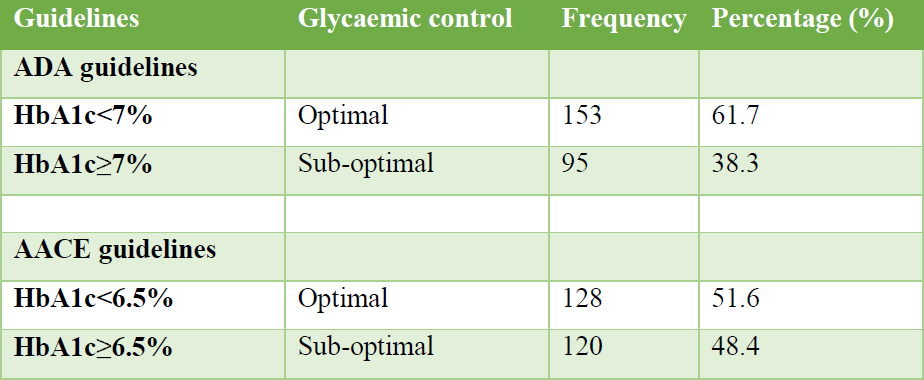
Glycaemic control of the participants

| Guidelines | Glycaemic control | Frequency | Percentage (%) |
| --- | --- | --- | --- |
| **ADA guidelines** |  |  |  |
| **HbA1c<7%** | Optimal | 153 | 61.7 |
| **HbA1c≥7%** | Sub-optimal | 95 | 38.3 |
| **AACE guidelines** |  |  |  |
| **HbA1c<6.5%** | Optimal | 128 | 51.6 |
| **HbA1c≥6.5%** | Sub-optimal | 120 | 48.4 |

**Table 5:** Differences in the means of certain parameters across the antidiabetic drugs classes.

| Antidiabetic drug | HbA1c |  | BMI |  | SBP |  | DBP |  |
| --- | --- | --- | --- | --- | --- | --- | --- | --- |
|  | T | p | T | p | T | p | T | p |
| Biguanide | 2.28 | 0.023** | -0.13 | 0.9 | -0.04 | 0.97 | 2.09 | 0.038** |
| DPP-IV inhibitors | 3.79 | 0.00** | 0.43 | 0.67 | 1.48 | 0.14 | 1.1 | 0.27 |
| SGLT-2 inhibitors | 0.83 | 0.41 | 0.54 | 0.59 | -0.16 | 0.87 | 0.37 | 0.57 |
| Insulin | 9.53 | 0.00** | -0.1 | 0.32 | 0.8 | 0.42 | 1.48 | 0.14 |
| GLP-1 agonists | -0.12 | 0.9 | 1.65 | 0.1 | 1.21 | 0.23 | 1.46 | 0.15 |
| Sulphonylureas | -0.54 | 0.59 | -1.16 | 0.25 | -0.29 | 0.77 | -0.6 | 0.55 |
| Thiazolidinediones | 1.05 | 0.3 | 1.49 | 0.14 | -0.77 | 0.44 | -<br>1.15 | 0.25 |
| α-glucosidase inhibitor | 0.44 | 0.66 | -0.11 | 0.92 | 0.93 | 0.35 | 0.41 | 0.68 |

\*\* - statistically significant
T- t-statistic
p- p-value
HbA1c- glycated haemoglobin
BMI- body mass index
SBP- systolic blood pressure
DBP- diastolic blood pressure

## Discussion

The study was conducted to determine the pattern of antidiabetic drugs used by patients with type 2 diabetes and assess their degree of glycaemic control. It was found that biguanides (85.5%) and DPP-IV inhibitors (60.9%) constituted the most commonly prescribed antidiabetic drugs. About 63% of the participants took 2-3 antidiabetic drugs. Only one-fifth of the participants were on monotherapy, which was mostly a biguanide. The mean HbA1c of the participants was 7.2±1.99 %. Using the ADA guideline, about two-thirds of the participants had optimal glycaemic control. Glycaemic control correlated significantly with the number of antidiabetic drugs used but was not significantly related to the modality of payment, whether out-of-pocket or via health insurance schemes.

The average age of the patients was 59.6 ± 12.7 years. Several studies done in Nigeria have reported that type 2 diabetes is most common in middle age of life, which is consistent with the findings of the present study.^9–11^ Insulin resistance increases with age and other risk factors for type 2 diabetes such as obesity and hypertension tend to peak at the same period.^12^ The study showed a slight male preponderance (57.7%). It has been previously documented that diabetes is more common among men.^13, 14^ Men have larger visceral fat mass and this is one of the plausible explanations for the observation.^15^ However, Adewumi et al^11^, in a study done in Nigeria, found equal prevalence across both genders while Chukwu et al^16^ even documented a higher female preponderance. The differences in study populations and designs could partly explain these discrepancies.

The mean BMI of the patients was 32.4±7.4 kg/m^2^ and less than 20% of the participants had a normal weight. Some studies in the past have quoted a frequency of normal weight among individuals living with type 2 diabetes as 15-21%.^17–19^ However, a few studies done in Nigeria and Uganda have quoted a frequency of normal weight as high as 37-41%. This difference in the proportion of individuals with normal weight may be due to the differences in study settings and methodology. Interestingly, all the studies with higher frequencies of normal weight were carried out in public settings typically attended by individuals in the medium and low socioeconomic classes whereas the present study was done in a private setting usually patronized by individuals of high socioeconomic status.

The present study showed that 48.4% of the participants exclusively paid for their care out of pocket while 51.6% had some degree of insurance coverage. The present study was carried out in a private tertiary hospital in an urban area in Nigeria. This finding is in total disagreement with what is obtainable in public institutions in Nigeria where as high as 85-100% of diabetic patients pay exclusively out of pocket to access care.^20, 21^ However, in the developed world, the reverse is the case, where about 85-100% of the patients with diabetes have health insurance coverage.^22–24^. So, the present study shows a payment pattern in-between that is typically seen in developing and developed countries.

Biguanides (85.5%) and DPP-IV inhibitors (60.9%) were the most commonly used antidiabetic drugs while thiazolidinediones (1.2%), α-glucosidase inhibitors (0.8%), and meglitinides (0%) were the least prescribed antidiabetic medications. In addition, 31.0% of the patients were on SGLT-2 inhibitors, 17.1% were on insulins, 15.3 were on GLP-1 agonists, and 14.9% were on sulphonylureas. Some studies done in Nigeria have equally reported that biguanides are the most commonly prescribed antidiabetic drugs (79.1-100.0%).^25–27^ Metformin is highly efficacious, cheap, safe and readily available. Most clinical practice guidelines still recommend metformin as the first-choice antidiabetic agent. However, when compared with previous studies in Nigeria, DPP-IV inhibitors were more frequently used in the present study (60% vs 0-6%).^25, 28^

Additionally, the cardioprotective GLP-1 agonists and SGLT-2 inhibitors were hardly used in the previous studies done in Nigeria.^25–28^ The present study was done in a setting where patients can directly or indirectly (through high-premium health insurance) afford these medications. This is further corroborated by studies done in advanced countries (France and Austria) which clearly demonstrated a high frequency of prescription of DPP-IV inhibitors, GLP-1 agonists and SGLT-2 inhibitors.^29, 30^ Previous studies have demonstrated comparable insulin usage (10.5-13.3%)^25, 28, 31^ with was found in this study (17.1%). The clinical practice guidelines recommend the commencement of insulins, especially basal insulins, in certain instances, one of which being poorly controlled type 2 diabetes, evidenced by markedly elevated HbA1c.

Averagely, about 2-3 drugs were prescribed to individuals with type 2 diabetes, as shown in the current study. This is consistent with prior studies which have reported a similar finding.^25, 27, 28^ Type 2 diabetes has different mechanisms underlying its pathogenesis and using medications targeting different pathways has been shown to improve glycaemic control and minimize the risk or slow down the progress of complications.^32, 33^ This study also found a statistically significant relationship between the number of antidiabetic drugs used and the age groups (Pearson Chi-square =45.06, p=0.038). In other words, the older age groups tend to use more drugs. In a study done in South-East Nigeria, Kennedy et al., did report that the older age groups needed more than monotherapy to enhance their glycaemic control.^28^ This could be due to the lifetime progressive decline in β-cell mass with a resultant effect in worsened dysglycaemia, and more drugs would be needed to counteract this progression.^34^ Simultaneously, due to increasing visceral fat mass, insulin resistance worsens and the frequencies of co-morbidities such as dyslipidaemia and hypertension rise as well and all these would warrant the use of multiple antidiabetic agents.

However, the number of antidiabetic drugs had no statistically significant association with BMI. This probably implies that polypharmacy was not necessarily associated with weight gain. This is possible because 4 (biguanide, DPP-IV inhibitors, SGLT-2 inhibitors and GLP-1 agonists) out of the top 5 most frequently used drugs in this study are either weight neutral or promote outright weight loss. Most of the studies done in Nigeria have not tested for a possible association between antidiabetic polypharmacy and weight changes. Interestingly, the number of drugs being used was not dependent on the modalities of payment (out-of-pocket vs insurance). Ordinarily, one would have taught that individuals paying out of pocket would be prescribed fewer drugs due to cost but this was not the case in this study. This is because in the private setting, affordability is often not an issue but the appropriateness and the results of the therapy.

Using an HbA1c cut-off of 7% (according to the ADA guidelines), 61.7% of the participants had optimal glycaemic control. This may be considered low but it is of interest to note that most studies in Nigeria have reported a much lower proportion of type 2 diabetes patients with optimal glycaemic control (16-46%).^35–38^ However, a few studies reported a much closer frequency of optimal glycaemic control (54-60%) but still slightly lower than the 61.7% seen in the current study.^39, 40^ This clearly shows that the proportion of individuals with optimal glycaemic control in this study is relatively substantial compared with prior studies. This could be due to the differences in study design and participants but it could also be due to the variations in the types of medications often used, as earlier highlighted. Furthermore, the current study did find a statistically significant negative correlation between the number of drugs used and HbA1c. Additionally, the use of metformin and DPP-IV inhibitors was associated with a significantly lower HbA1c. Previous studies have demonstrated the efficacy of metformin and DPP-IV inhibitors, especially when combined (which was a common observation in this study).^41–43^ This may be partly due to their high tolerability, low risk of hypoglycaemia, simplicity of administration and ultimately enhanced adherence.^44^

### Strengths of the study

The study was carried out in a developing country but a high-profile private setting where there is no scarcity of resources. It is an opportunity to demonstrate what could happen if there are better resources in the nation or if the health sector is better structured. It shows a clear departure from the old pattern, where sulphonylureas without cardioprotective properties were more frequently prescribed. Now, modern antidiabetic medications with good cardiovascular safety are more frequently prescribed especially in a private setting, as seen in this study. The study has demonstrated how this paradigm shift can potentially lead to improved glycaemic control.

### Limitations of the study

The study only implied strong adherence to the prescribed medications but this was not confirmed with any drug assay.

## Conclusions

The pattern of antidiabetic drugs has evolved, especially in a private setting where affordability is not a concern. The present trend favours the use of costlier but safer, more cardiovascular-friendly and possibly more efficacious drugs such as DPP-IV inhibitors, SGLT-2 inhibitors and GLP-1 agonists. Biguanides remain the drug of choice though. Combining 2 to 3 drugs is very common. Glycaemic control is better in the present study and it could be due to the pattern of drugs being used in this study. This study will serve as a pointer to how pattern of drugs could affect HbA1c even in a developing country. Policymakers could also use this study as a justification for reforms in the public health sector. It could also be a starting point for bigger studies in the future

## List of abbreviations

AACE/ACE: American Association of Clinical Endocrinologists/American College of Endocrinology
ADA: American Diabetic Association
BMI: Body mass index
DBP: Diastolic blood pressure
DPP-IV: Dipeptidyl peptidase-IV
GLP-1: Glucagon-like peptide-1
HbA1c: Glycated haemoglobin
PPAR-□: Peroxisome proliferator-activated receptor-gamma
SBP: Systolic blood pressure
SGLT-2: Sodium-glucose co-transporter 2
WHO: World Health Organization

## Declarations

### Acknowledgement

None

## Author contributions

### Ethical approval and consent to participate

Obtained from the Ethical Committee of the hospital

### Funding

No external funding was received

### Availability of data

Available on request

## Data Availability

All the data and materials are available on reasonable request

